# Himalayan Tartary Buckwheat Sprout Powder, Perceived Cognitive Abilities, and Objective Processing Speed: A Randomized, Double-Blind, Placebo-Controlled Trial

**DOI:** 10.64898/2026.11.03.26367186

**Authors:** Kaitlyn P. White, Jaime L. Tartar, Macarena Roel, Varun B. Dwaraka, Minh Chau, Austin Perlmutter, Valerie Starratt, Jeff Chen, Susan J. Hewlings

**Author notes:** Correspondence: Kaitlyn P. White, MA, MS Radicle Science, Inc.

## Abstract

Subjective cognitive difficulties are common in midlife and later life, and flavonoid-rich foods may support everyday cognitive function. Tartary buckwheat (*Fagopyrum tataricum*) sprouts are rich in rutin and other flavonoids, but their cognitive effects have not been tested in humans. In this randomized, double-blind, placebo-controlled trial, US adults with self-reported cognitive difficulties received 18 g per day of Himalayan Tartary buckwheat (HTB) sprout powder or matched placebo for 12 weeks. Of 848 randomized participants, 662 were analyzed. While the rate of change in the primary outcome, PROMIS Cognitive Function, did not differ between arms (contrast = 0.03, 95% CI −0.07 to 0.13; p = .59), PROMIS Cognitive Abilities improved nominally faster with HTB (contrast = 0.12, 95% CI 0.00 to 0.24; p = .046), but not after Bonferroni adjustment. Among participants with final-week data (n = 296), more HTB than placebo participants achieved a meaningful (≥0.5 SD) improvement in Cognitive Abilities (68.7% vs 52.5%; RR = 1.32, 95% CI 1.10 to 1.59; p = .003). Fatigue, sleep-related impairment and depressive symptoms did not differ between arms. In exploratory computerized testing (n = 512), HTB participants had faster reaction times on the Flanker and Stroop tasks, without loss of accuracy. Exploratory epigenetic analyses (n = 34) identified hypothesis-generating signals in epigenetic age and immune-cell composition, none surviving multiple-testing correction. Side-effect rates were similar between arms. HTB sprout powder did not improve the primary outcome, but findings for perceived cognitive abilities and processing speed warrant confirmation in adequately powered trials. Trial registration: NCT07245784.

## Introduction

Self-perceived cognitive difficulties are common in midlife and later life and are an important public health concern. In 2023, 16.9% of US adults aged 45 years or older reported subjective cognitive decline (confusion or memory loss that was happening more often or getting worse), and 59% of those affected were worried about it.¹ Beyond this day-to-day burden, subjective cognitive complaints are also associated with an approximately twofold higher risk of subsequent mild cognitive impairment and dementia.²˒³ Because these complaints can impose a meaningful day-to-day burden and may also signal early decline, safe and accessible strategies that help adults maintain everyday cognitive function are of considerable interest.

Diet is a modifiable factor that may influence cognitive health, and dietary flavonoids have received particular attention. Flavonoids and other polyphenols may support brain health through anti-inflammatory, antioxidant, metabolic, vascular, gut microbiome and epigenetic pathways.⁴ In two large prospective US cohorts, higher long-term intake of total flavonoids was associated with lower odds of subjective cognitive decline.^5^ In a large prospective cohort, higher intake of flavonoid-rich foods was also associated with lower dementia risk,^6^ and in a meta-analysis of 80 randomized controlled trials, flavonoid interventions produced a small overall improvement in cognitive performance (Hedges’ *g* = 0.15), with significant benefits in chronic interventions and in middle-aged and older adults.^7^ However, these trials were generally small (5519 participants across 80 studies) and focused largely on performance on cognitive tests rather than on how people perceive their own cognition. Whether flavonoid-rich whole foods improve how adults perceive their everyday cognitive function, and whether any such change is mirrored in objective measures of attention and processing speed, therefore remains unclear.

Controlled food-based interventions further support the cognitive and epigenetic effects of polyphenol-rich diets. In a 3-month randomized, double-blind, placebo-controlled trial in middle-aged adults, Montmorency tart cherry supplementation improved sustained attention and reduced self-reported mental fatigue relative to placebo.^8^ Similarly, in a randomized, double-blind, placebo-controlled crossover trial, a polyphenol-rich fruit, vegetable and berry juice powder concentrate improved cognitive performance relative to placebo and increased plasma CREB and BDNF, suggesting potential benefit to neuroplasticity.^9^ Polyphenols can also modulate gene regulation through epigenetic mechanisms, including DNA methylation, histone modifications, and microRNA expression.^10^ Consistent with this possibility in humans, the 18-month DIRECT-PLUS randomized trial found that a polyphenol-rich green Mediterranean diet produced distinct blood DNA methylation and gene-expression profiles relative to comparator diets.^11^ A separate analysis of the same trial found attenuated age-related brain atrophy, most prominently with the high-polyphenol diet.^12^ These findings provide a rationale for evaluating both cognitive outcomes and epigenetic responses in trials of polyphenol-rich foods.

One food of interest in this context is Tartary buckwheat (*Fagopyrum tataricum* (L.) Gaertn.), a pseudocereal that originated in the Himalayan region and has long been cultivated as a food and medicinal crop.^13, 14^ Comparative analyses have found higher rutin and total phenolic acid contents in Tartary buckwheat seeds than in common buckwheat (*Fagopyrum esculentum* Moench),^15^ alongside other flavonoids, D-chiro-inositol, and bioactive peptides and polysaccharides.^16^ Sprouting further shapes this profile. During germination, total flavonoid concentrations can increase up to threefold over the first four days.^17,18^ Sprouts retain high rutin levels after boiling and steaming, making them a practical dietary source of these compounds.^19^ Sprouting may also improve protein digestibility and increase certain essential amino acids, γ-aminobutyric acid (GABA) and D-chiro-inositol.^20, 21^

Human studies of Tartary buckwheat have focused mainly on cardiometabolic and some immune outcomes. In an open-label randomized trial in 165 adults with type 2 diabetes, replacing part of the daily staple food with Tartary buckwheat for 4 weeks reduced fasting insulin, total cholesterol, and low-density lipoprotein cholesterol relative to a control diet.^22^ In a double-blind crossover trial comparing Tartary and common buckwheat cookies, serum myeloperoxidase, an inflammatory marker, decreased during the Tartary buckwheat period.^23^ Evidence from broader buckwheat studies is less consistent. A 2018 systematic review and meta-analysis reported reductions in blood glucose, total cholesterol and triglycerides with buckwheat consumption.^24^ However, that analysis pooled cross-sectional and intervention data. A later meta-analysis that excluded cross-sectional studies and combined different types of buckwheat found only small, non-significant effects, and most of its included studies raised concerns about risk of bias.^25^ Because the nutrient profiles of common and Tartary buckwheat differ substantially, combining multiple forms of this crop into pooled research could obscure the potential effects of Tartary buckwheat when studied as a standalone intervention.^15^ Beyond cardiometabolic outcomes, a 90-day single-arm, open-label pilot study in 50 healthy adults reported that a polyphenol-rich supplement largely based on the phytochemical composition of Tartary buckwheat was associated with changes in epigenetic age clocks and immune markers.^26^ These human data indicate a potential for polyphenol– and especially flavonoid-rich Tartary buckwheat to influence metabolic and inflammatory markers, pathways also implicated in brain health.^4^ Evidence linking Tartary buckwheat directly to brain function, however, remains preclinical. Tartary buckwheat extracts improved cognition and memory in mice with amyloid-β-induced impairment,^27^ attenuated neurobehavioral impairments in ovariectomized rats,^28^ reduced acrylamide-induced neurotoxicity in zebrafish larvae,^29^ and reduced fatigue-like behavior in a mouse model of central fatigue.^30^ In senescence-accelerated mice, however, dietary common buckwheat whole flour attenuated age-related cognitive decline, whereas Tartary buckwheat whole flour did not.^31^ To our knowledge, no published randomized controlled trial has evaluated Tartary buckwheat or its sprouts for cognitive outcomes in humans.

We therefore conducted a decentralized, randomized, double-blind, placebo-controlled trial of daily Himalayan Tartary buckwheat sprout powder (18 g per day, mixed into a beverage) in US adults selected for potential improvement in self-reported cognitive function across a 12-week intervention period. Our aim was to determine whether the sprout powder improves self-perceived cognitive function relative to placebo, and whether any change extends to related patient-reported outcomes, computerized measures of attention and inhibitory control, and epigenetic biomarkers.

## Methods

### Study Design

Over a 12-week intervention period, this decentralized, randomized, double-blind, placebo-controlled, parallel-group trial evaluated the safety and effects of Himalayan Tartary Buckwheat Sprout Powder on cognitive function and related health outcomes in adults residing in the United States. The study was conducted entirely through a direct-to-consumer virtual platform, with no in-person study visits. Participants self-administered either the active study product or matched placebo at home and completed all study assessments electronically through survey links delivered by text message or email.

### Trial Registration and Oversight

The trial was registered at ClinicalTrials.gov (NCT07245784) under protocol number RADX_P_2406_RBB and was approved by the Sterling Institutional Review Board (approval number 14726-SHewlings). It was conducted in accordance with the Declaration of Helsinki, the Belmont Report and applicable US regulations (21 CFR Parts 50, 54 and 56; 45 CFR Part 46), and all participants provided electronic informed consent. Recruitment ran from 4 December 2025 to 2 February 2026, and the study ended on 19 May 2026.

### Participants

Eligible participants were adults aged 21 years or older who resided in the United States, could read and understand English, and were willing to use an unidentified study product for 12 weeks without knowing whether they had been assigned to active treatment or placebo. Participants were also required to have the potential for at least 30% improvement in their primary health outcome (assessed with the PROMIS Cognitive Function 4a at screening), provide a valid U.S. shipping address and mobile telephone number, and have reliable daily internet access. Screening also included a reading-comprehension item and consent-comprehension questions.

Individuals were excluded if they were pregnant, attempting to become pregnant, or breastfeeding; were concurrently enrolled in another clinical trial; reported heavy alcohol consumption, defined as three or more alcoholic beverages per day; reported a documented food allergy to milk, soy, egg, wheat, or nuts; or reported a recent major illness or surgery that presented a significant safety concern. Additional exclusions included cardiac, hepatic, or renal conditions associated with a known contraindication to study ingredients, including advanced heart failure, atrial fibrillation, cirrhosis, advanced chronic kidney disease, or kidney failure.

Participants were also excluded for use of medications with an established moderate-to-severe interaction with study-product ingredients, including selected anticoagulants, antihypertensives, psychotropic medications, thyroid medications, diabetic medications, and other specified therapies. Current chemotherapy or immunotherapy, and current use of supplements containing buckwheat sprout powder, polyphenols or flavonoids (or similar products that could interfere with assessment of the intervention), were also exclusionary.

Participants were recruited remotely through internet-based networks and email.

### Randomization

Participants who completed baseline assessments were stratified by age, sex assigned at birth, and baseline health outcome score before 1:1 randomization to maintain balance across study arms. Participation in the optional biospecimen sub-study was included as an additional stratification factor when applicable. Assignments were double-blinded until the trial’s conclusion.

### Study Products

Both products were supplied as powder stick packs. The active product (RBB31) contained 18,000 mg of Himalayan Tartary buckwheat sprout powder (HTB). The placebo (RBB30) was a matched powder made with rice powder and natural caramel color. Participants were instructed to mix one stick pack into at least 6 oz of cold water, milk, a milk alternative, juice or another beverage once daily for 12 weeks, consistent with the study’s real-world-use design.

### Study Procedures

Participants first completed electronic informed consent through a secure online platform. Brief comprehension questions were embedded throughout the consent form, and participants were required to answer them correctly before proceeding. Participants could contact study personnel by telephone, email, or online chat if clarification was needed. After providing consent, participants verified their identity using a secure electronic identity-verification service and completed eligibility screening, demographic and intake questionnaires, address verification, and baseline assessments.

After randomization, a 12-week supply of study products was then shipped directly to each participant’s home along with instructions for use and study participation. During the 12-week intervention period, participants completed an initial assessment regarding their impression of the product and any effects noticed after the first serving. Weekly electronic assessments were subsequently administered to collect information regarding study-product use, side effects, satisfaction, and health outcomes. Randomly positioned attention-check questions were embedded in the baseline and weekly surveys to support data integrity and identify inattentive responding.

Participants could additionally elect to participate in application-based cognitive testing and/or at-home biospecimen collection. Participants who opted into computerized cognitive testing completed cognitive assessments at baseline and during follow-up. Participants enrolled in the biospecimen sub-study provided samples before and after study-product use for exploratory biomarker analyses.

At study completion, participants completed a final survey assessing their overall experience and perceived effects of the study product. Participants were then unblinded to treatment assignment and received a personalized descriptive report summarizing the health information they had recorded during the study.

### Outcome Measures

The primary outcome was PROMIS Cognitive Function. Powered secondary outcomes were PROMIS Fatigue, PROMIS Sleep-Related Impairment, and PROMIS Cognitive Abilities assessments. Exploratory outcome measures included PROMIS Emotional Distress–Depression Short Form, computerized cognitive outcomes, and DNA methylation–based measures of biological aging and immune-cell composition.

#### Cognitive Function

The primary outcome was perceived cognitive function, assessed using the Patient-Reported Outcomes Measurement Information System (PROMIS) Cognitive Function Short Form 8a. This measure evaluates a participant’s perception of cognitive difficulties experienced in everyday life, including problems with concentration, memory, mental clarity, processing information, and performing cognitively demanding activities. The PROMIS Cognitive Function short forms demonstrated high internal consistency (Cronbach’s α = .85–.95).^32^ Higher scores indicate better perceived cognitive function. Scores were converted to T-scores, and baseline severity categories (within normal limits, mild, moderate, severe) were assigned using PROMIS T-score thresholds.

#### Fatigue

Fatigue was a powered secondary outcome assessed using the PROMIS Fatigue Short Form 8a. PROMIS Fatigue measures the experience and impact of fatigue, including feelings of tiredness, exhaustion, reduced energy, and the extent to which fatigue interferes with daily functioning. Higher scores indicate greater fatigue. The PROMIS Fatigue 8a demonstrated excellent internal consistency (Cronbach’s α ≥ .90) and test-retest reliability (intraclass correlation [ICC] ≥ .90).^33,34^

#### Sleep-Related Impairment

Sleep-related impairment was a powered secondary outcome assessed using the PROMIS Sleep-Related Impairment measure. This domain assesses perceived functional consequences of sleep problems during waking hours, including sleepiness, difficulty remaining alert, impaired concentration, irritability, and reduced ability to function because of poor sleep. The PROMIS Sleep-Related Impairment 8-item short form demonstrated convergent (*r* = .68) and discriminant (*r* = .46) validity and differentiated individuals with and without self-reported sleep disorders.^35^

#### Cognitive Abilities

Perceived cognitive abilities were assessed as a powered secondary outcome using the PROMIS Cognitive Function–Abilities Subset. It assesses a participant’s perceived capacity to perform cognitive activities. These include mental sharpness, memory, concentration, clear and efficient thinking, attention, and the ability to keep track of tasks despite interruption. The PROMIS Cognitive Abilities demonstrated strong internal consistency (α = .85–.95) and construct validity across general and clinical populations.^36–38^

#### Mood

Depressive symptoms were evaluated as an exploratory outcome using the PROMIS Emotional Distress–Depression Short Form 8a. The PROMIS Depression measure assesses negative mood, views of the self, social cognition, and reduced positive affect over the preceding 7 days. Higher scores indicate greater depressive symptom burden. The PROMIS Depression 8a demonstrated excellent internal consistency (α = .95).^39^

#### Computerized Cognitive Outcomes

Participants who opted into the application-based cognitive testing completed objective computer-based cognitive assessments using Inquisit Web (version 7.1.5; Millisecond Software, LLC, Seattle, WA, USA). The battery included the Flanker task, Stroop task, and Sustained Attention to Response Task (SART).

The Flanker task assessed selective attention and interference suppression. Outcomes included mean reaction time for correct responses and interference reaction time; lower reaction times indicate faster performance.

The Stroop task assessed selective attention and inhibitory control by requiring participants to respond to relevant stimuli while suppressing competing information. Outcomes included mean correct reaction time and interference reaction time.

The SART assessed sustained attention and response control. Outcomes included sensitivity (*d′*), commission-error percentage, and the coefficient of variation of reaction time. Higher *d′* indicates better discrimination and sustained attention, whereas higher commission-error rates or greater reaction-time variability indicate poorer response control or consistency.

Accuracy was derived from task output for the Flanker, Stroop, and SART and examined alongside reaction time to assess potential speed–accuracy trade-offs. For the Stroop task, mean correct reaction time was also evaluated across the four congruency conditions.

#### Epigenetic Biomarkers

Participants in the optional biospecimen sub-study self-collected a blood sample at home at baseline and at the end of the 12-week intervention. DNA methylation profiling and computation of methylation-based measures were performed by TruDiagnostic (Lexington, KY, USA). Genome-wide DNA methylation was measured with a custom Illumina Infinium MethylationEPIC v2.0 array, and raw data were preprocessed in R with minfi using single-sample Noob normalization, with samples and probes filtered on detection p values.^40^

Four DNA methylation clocks were designated as the primary epigenetic outcomes because they had been evaluated in the earlier single-arm pilot study:^26^ OMICmAge,^41^ DunedinPACE,^42^ and principal-component versions of PhenoAge and GrimAge (PCPhenoAge and PCGrimAge).^43–45^ All other epigenetic clock, mitotic clock, and immune-cell deconvolution measures produced by the laboratory’s standard pipeline (370 measures) were analyzed as a single exploratory screen. Further exploratory analyses included an epigenome-wide association study, a targeted analysis of NF-κB pathway genes and KEGG pathway enrichment (Supplementary Table E5) and methylation-based surrogates of circulating proteins and metabolites.

#### Safety

Safety was evaluated using weekly participant reports of side effects. Participants completed a structured symptom checklist that included gastrointestinal symptoms, nausea, headache, dizziness, constipation, dry mouth, drowsiness, insomnia, and other potential adverse effects. Follow-up information could include symptom onset, severity, study-product use at the time of onset, and medical care sought. Side effects were summarized by study arm to characterize the frequency and overall safety profile of the intervention. Serious adverse events and unanticipated problems were reviewed by the principal investigator.

### Statistical Analysis

#### Sample Size and Power

Sample size was determined using a Monte Carlo power analysis based on 1,000 simulations. The analysis was designed to provide 80% power to detect a small between-group difference in rates of change at a two-sided α = .05. Allowing for up to 40% attrition, the target sample size was 250 participants per study arm. Participants who discontinued the study were not replaced.

#### Analysis Populations

The primary analysis population (a modified intent-to-treat population, hereafter ITT) included randomized participants who completed at least one postbaseline weekly assessment, analyzed by assigned arm. The per-protocol (PP) population, analyzed as an exploratory analysis, included participants with ≥80% dosing adherence who completed all required weekly assessments. The cognitive ITT (cogITT) cohort included participants who completed computerized cognitive testing at baseline and at least one postbaseline assessment.

Primary and powered secondary PROMIS outcomes were analyzed using Linear Mixed-Effects (LMM) models, adjusting for sex at birth, age, body mass index, baseline outcome value and the baseline × week interaction, with a random intercept and random slope for week (unstructured covariance) and robust standard errors. Models accounted for repeated measurements within participants and evaluated the treatment group × week interaction to determine whether change over time differed between treatment and placebo.

For each PROMIS outcome, meaningful improvement was defined as a change of at least 0.5 SD of the baseline score distribution in the direction of improvement between baseline and the final study week.^46^ Participants meeting this threshold were classified as meaningful responders. These analyses were restricted to participants who completed the final-week survey. The probability of achieving a meaningful improvement was compared between HTB and placebo using Poisson regression with robust error variance, adjusted for sex at birth, age, and body mass index, and reported as relative risks (RRs) with 95% confidence intervals.

Exploratory computerized cognitive outcomes were analyzed using the Mixed Model for Repeated Measures (MMRM), adjusting for baseline performance. Models evaluated treatment-group and treatment group × week effects for each cognitive outcome separately. Side effects were summarized by study arm and compared using Pearson’s χ² or Fisher’s exact tests, as appropriate.

Multiplicity was controlled using family-wise Bonferroni corrections. The primary outcome was tested at α = .05. Rate-of-change comparisons for the powered secondary outcomes (fatigue, sleep-related impairment and cognitive abilities) were evaluated at a Bonferroni-adjusted α = .0167. Meaningful responder analyses were corrected across all five PROMIS outcomes (α = .01). Rate-of-change analyses of depressive symptoms, per-protocol analyses, and computerized cognitive and epigenetic outcomes were exploratory, were not adjusted for multiplicity, and are interpreted as hypothesis-generating.

Epigenetic outcomes were analyzed in participants with usable baseline and end-of-intervention samples. For each measure, the between-arm difference in change from baseline was tested with a treatment × timepoint interaction in a linear mixed model adjusted for age and sex, with a random effect for participants. Effect sizes are reported as unadjusted Cohen’s d for the between-arm difference in change scores; negative values indicate a smaller increase or a larger decrease in the HTB arm than in the placebo arm. The Benjamini–Hochberg false discovery rate (FDR) was controlled separately within the four primary clocks and within the exploratory screen.^47^ With 15 HTB and 19 placebo participants, the sub-study had 80% power to detect only large between-arm differences (d ≥ 1.00, two-sided α = .05), and all epigenetic analyses were considered hypothesis-generating.

## Results

### Participant Flow and Characteristics

Of 848 randomized participants, 662 (348 placebo, 314 HTB) completed at least one postbaseline weekly survey and were included in the ITT analyses of the PROMIS outcomes. Weekly survey response rates ranged from 40% to 86% and were similar across arms. The per-protocol (PP) cohort included 125 participants (75 placebo, 50 HTB). Among participants who opted into computerized cognitive testing, 512 (256 per group) completed testing at baseline and at least one follow-up and comprised the cogITT cohort.

Baseline demographic and clinical characteristics are presented in Table 1. Among participants represented in the baseline table (*N* = 662), mean age was 48.72 years (*SD* = 11.34), 62.8% were female, and 77.0% were White. Most participants (82.3%) reported experiencing poor attention or focus for at least 3 months. Baseline PROMIS severity distributions were generally similar between study arms.

**Table 1.** Demographic and Baseline Characteristics of Study Participants.

| **Characteristic** | **Placebo** | **HTB** | **Total** |
|---|---|---|---|
| N (%) | 348 (52.6%) | 314 (47.4%) | 662 (100.0%) |
| Age, M ± SD (min–max) | 49.30 ± 11.19<br>(21–75) | 48.08 ± 11.49<br>(21–75) | 48.72 ± 11.34<br>(21–75) |
| BMI, M ± SD | 28.16 ± 6.83 | 28.21 ± 6.21 | 28.19 ± 6.54 |
| **BMI category** |  |  |  |
| Underweight | 11 (3.2%) | 6 (1.9%) | 17 (2.6%) |
| Normal weight | 117 (33.6%) | 99 (31.5%) | 216 (32.6%) |
| Overweight | 100 (28.7%) | 110 (35.0%) | 210 (31.7%) |
| Obesity | 120 (34.5%) | 99 (31.5%) | 219 (33.1%) |
| **Sex** |  |  |  |
| Male | 127 (36.5%) | 119 (37.9%) | 246 (37.2%) |
| Female | 221 (63.5%) | 195 (62.1%) | 416 (62.8%) |
| **Race** |  |  |  |
| White | 268 (77.0%) | 242 (77.1%) | 510 (77.0%) |
| Asian | 17 (4.9%) | 16 (5.1%) | 33 (5.0%) |
| Black | 15 (4.3%) | 14 (4.5%) | 29 (4.4%) |
| Hispanic or Latino | 14 (4.0%) | 11 (3.5%) | 25 (3.8%) |
| Middle Eastern or North African | 0 (0.0%) | 2 (0.6%) | 2 (0.3%) |
| Multi-racial | 31 (8.9%) | 26 (8.3%) | 57 (8.6%) |
| Native Hawaiian or Pacific Islander | 1 (0.3%) | 0 (0.0%) | 1 (0.2%) |
| Some other race | 2 (0.6%) | 2 (0.6%) | 4 (0.6%) |
| Prefer not to say | 0 (0.0%) | 1 (0.3%) | 1 (0.2%) |
| **Race (collapsed)** |  |  |  |
| White | 268 (77.0%) | 242 (77.1%) | 510 (77.0%) |
| Non-white | 80 (23.0%) | 71 (22.6%) | 151 (22.8%) |
| Prefer not to say | 0 (0.0%) | 1 (0.3%) | 1 (0.2%) |
| **Experiencing poor attention/focus for 3+ months** |  |  |  |
| No | 62 (17.8%) | 55 (17.5%) | 117 (17.7%) |
| Yes | 286 (82.2%) | 259 (82.5%) | 545 (82.3%) |

*Note.* BMI = Body Mass Index

### Safety

During the 12-week intervention, the proportion of participants reporting side effects did not differ significantly between the HTB (18.8%) and placebo (15.2%) groups (*p* = .22). The most commonly reported side effects were gastrointestinal, including upset stomach or cramping, diarrhea, and nausea. Side effects were mostly mild in both arms. One serious adverse event, which required emergency healthcare services, was reported in the HTB arm and assessed as possibly related to the study product; a causal relationship could not be confirmed or excluded.

### Primary Outcome

In the ITT cohort, LMM analysis showed that the rate of change in PROMIS Cognitive Function did not differ significantly between treatment and placebo (estimated weekly slopes 0.26 [95% CI 0.18, 0.34] for HTB and 0.23 [0.17, 0.30] for placebo; contrast = 0.03, SE = 0.05, 95% CI [−0.07, 0.13], *p* = .59) (Figure 1).

**Figure 1.**
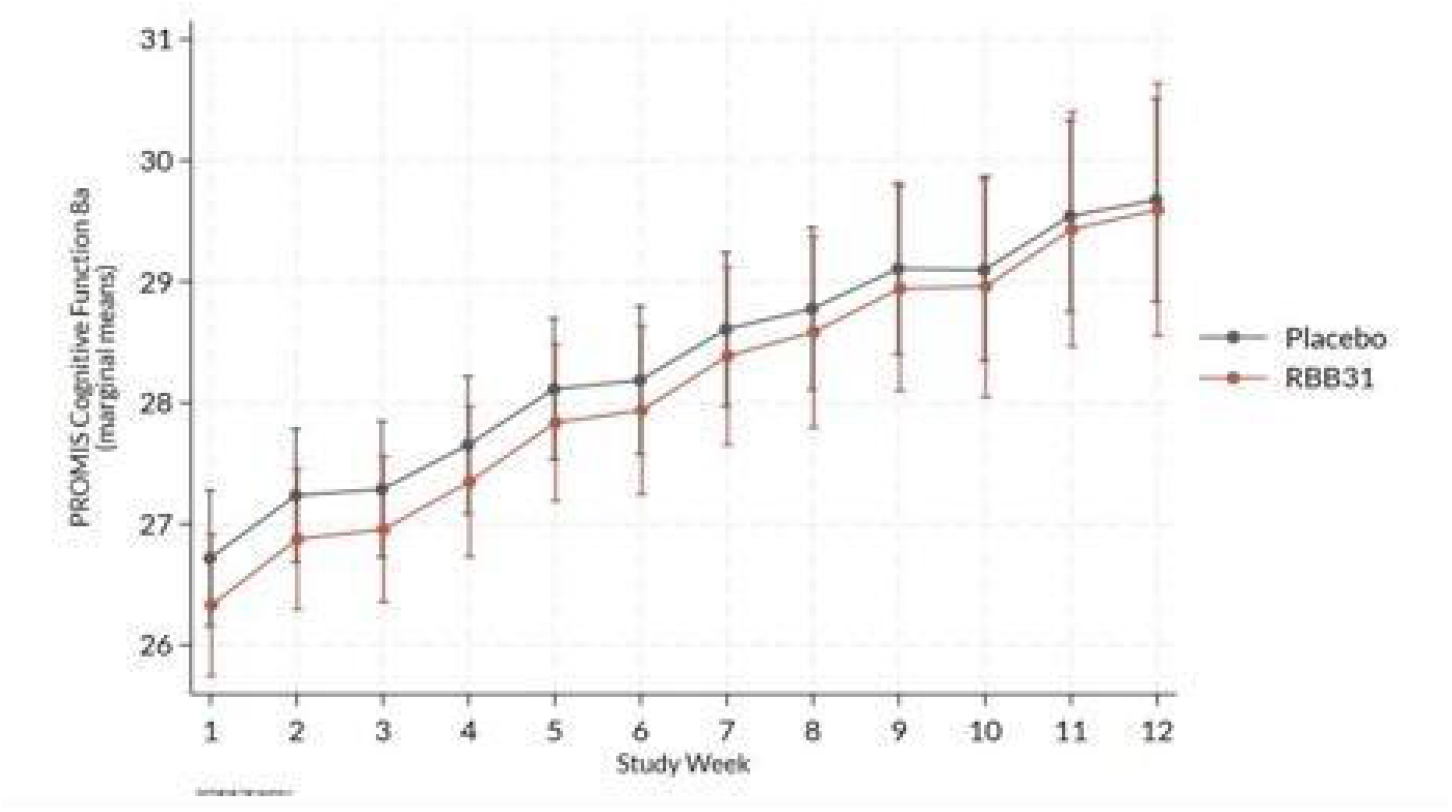
LMM-Estimated PROMIS Cognitive Function Scores Over Time by Study Arm.

### Secondary and Exploratory PROMIS Outcomes

For Cognitive Abilities, LMM analysis demonstrated that the HTB group showed a nominally greater rate of improvement than placebo (contrast = 0.12, SE = 0.06, 95% CI [0.00, 0.24], p = .046). The estimated slopes were .27 (95% CI [.18, .36]) for treatment and .15 (95% CI [.07, 0.23]) for placebo. However, the between-group difference did not meet the Bonferroni-adjusted significance threshold of *α* = .0167 (Figure 2).

**Figure 2.**
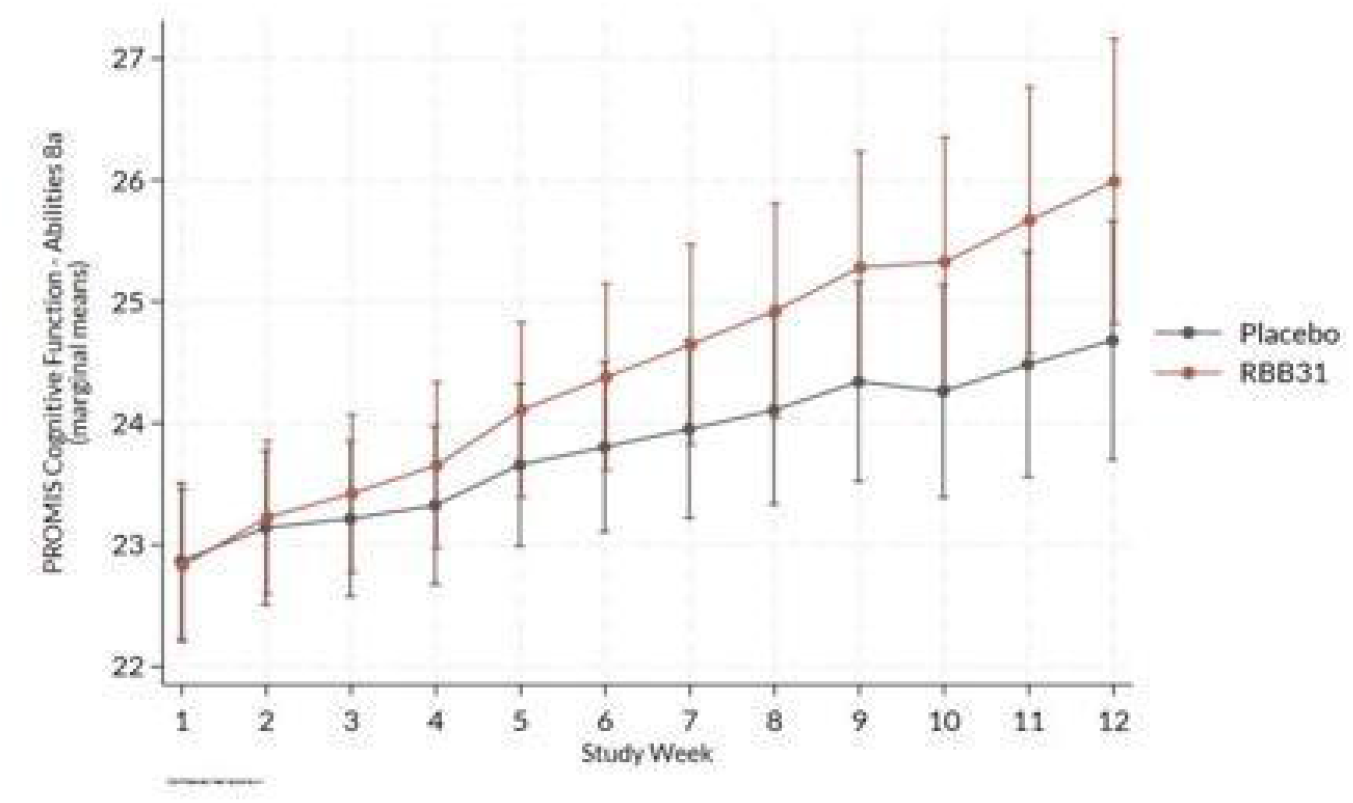
LMM-Estimated PROMIS Cognitive Abilities Scores Over Time by Study Arm. No significant between-group differences in rate of change were observed for PROMIS Fatigue, PROMIS Sleep-Related Impairment, or PROMIS Depression (p = .058, .122 and .189, respectively).

### Responder Analyses

Among the 296 participants with final-week data (162 placebo, 134 HTB), HTB participants (68.7%) were significantly more likely than placebo participants (52.5%) to achieve a meaningful improvement in Cognitive Abilities (RR = 1.32, 95% CI [1.10, 1.59], *p* = .003, below the Bonferroni-adjusted α = .01), corresponding to a 32% greater likelihood of meaningful improvement. The likelihood of achieving a meaningful improvement did not differ significantly between HTB and placebo for PROMIS Cognitive Function, Fatigue, Sleep-Related Impairment or Depression (all p ≥ .11).

### Per-Protocol Analyses (Exploratory)

In the PP cohort, the rate of change in PROMIS Cognitive Function did not differ significantly between treatment and placebo (contrast = 0.09, *SE* = 0.10, *p* = .37). HTB participants demonstrated greater rates of improvement than placebo participants in PROMIS Fatigue (contrast = −0.19, *SE* = 0.08, *p* = .02), with estimated slopes of −0.25 (95% CI [−0.34, −0.16]) for treatment and −0.06 (95% CI [−0.19, 0.06]) for placebo, and PROMIS Sleep-Related Impairment (contrast = −0.14, *SE* = 0.07, *p* = .03), with estimated slopes of −0.24 (95% CI [−0.33, −0.15]) and −0.09 (95% CI [−0.19, 0.00]), respectively. A greater rate of improvement was also observed for PROMIS Cognitive Abilities (contrast = 0.22, *SE* = 0.11, *p* = .046), with estimated slopes of 0.41 (95% CI [0.24, 0.58]) for treatment and 0.19 (95% CI [0.07, 0.31]) for placebo. The rate of change in PROMIS Depression did not differ significantly between groups (contrast = −0.09, *SE* = 0.06, *p* = .11).

### Exploratory Computerized Cognitive Outcomes

MMRM analyses showed that the treatment group demonstrated faster mean correct reaction times than placebo on the Flanker (estimate = −317.26 ms, *SE* = 83.47, 95% CI [-481.31, –153.22], *p* < .001) and Stroop tasks (estimate = −268.11 ms, *SE* = 67.57, 95% CI [-400.92, –135.67], *p* < .001). A significant treatment group × week interaction was observed for Flanker mean correct reaction time (estimate = 37.12 ms, *SE* = 9.13, 95% CI [19.21, 55.02], *p* < .001), indicating that the difference in reaction time changed over the study period. There was no significant difference in interference reaction time for the Flanker or Stroop task (*p* > .05). For the SART, a significant treatment group × week interaction was observed for reaction-time variability (estimate = .003, *SE* = .001, 95% CI [.001, .005], *p* = .013), with a smaller reduction in variability in the HTB arm. Treatment and placebo did not differ significantly in sensitivity, commission errors, or overall reaction-time variability, and no significant interactions were observed for sensitivity or commission errors (*p* > .05).

To assess possible speed–accuracy trade-offs, overall accuracy was examined for each task. Accuracy did not differ between HTB and placebo on the Flanker, Stroop, or SART (all p ≥ .447). Treatment group × week interactions were also nonsignificant for all three tasks. In condition-specific analyses of Stroop reaction time, HTB participants responded faster than placebo across all four congruency conditions (all p ≤ .020), with no significant treatment group × week interactions (all p ≥ .116).

### Exploratory Epigenetic Biomarkers

Thirty-five sub-study participants provided paired baseline and end-of-intervention samples (15 HTB, 20 placebo). One placebo baseline sample failed quality control, leaving 34 participants (15 HTB, 19 placebo) for analysis (Supplementary Table E1). None of the four primary clocks differed between arms in change from baseline (OMICmAge, p = .24, d = 0.40; PCGrimAge, p = .44, d = 0.26; DunedinPACE, p = .63, d = 0.16; PCPhenoAge, p = .72, d = −0.12; Supplementary Table E2).

In the exploratory screen, 9 of 370 measures reached nominal significance (p < .05), fewer than the approximately 18 expected by chance, and none remained significant after FDR correction. The three largest between-arm differences were all relative decreases in the HTB arm: the OmniAge Pipek elastic-net epigenetic age deviation (DeltaAge; p = .009, d = −0.92), the estimated fraction of activated natural killer (NK) cells (p = .011, d = −0.89), and age-adjusted epiTOC2 mitotic age, an estimate of cumulative stem-cell divisions (p = .023, d = −0.79; Supplementary Table E3).^48^ In the epigenome-wide analysis, 534 CpGs reached nominal significance (p < 0.001) (Supplementary Table E4). In these further exploratory analyses, no CpG site, NF-κB pathway gene, or methylation-based surrogate showed a between-arm difference robust to multiple-testing correction and adjustment for immune-cell composition; the most consistent gene-level signal was at three CpG sites annotated to NFKB2 (Supplementary Tables E4-E6).

## Discussion

To our knowledge, this is the first randomized controlled trial of Tartary buckwheat for cognitive outcomes in humans. In this 12-week decentralized, randomized, double-blind, placebo-controlled trial, daily consumption of Himalayan Tartary buckwheat (HTB) sprout powder was associated with potential cognitive benefits relative to placebo, although it did not significantly alter the prespecified primary outcome of Cognitive Function. For the secondary outcome of Cognitive Abilities, participants receiving HTB showed a greater rate of improvement than participants receiving placebo, but the between-group difference did not meet the Bonferroni-adjusted significance threshold. In the responder analysis, 68.7% of HTB participants compared with 52.5% of placebo participants met the prespecified 0.5-SD threshold for improvement in Cognitive Abilities, a difference that remained significant after correction for multiple comparisons. In exploratory computerized testing, HTB participants also had faster reaction times on the Flanker and Stroop tasks without loss of accuracy. No between-group differences were observed in fatigue, sleep-related impairment, or depressive symptoms in the longitudinal ITT population. In exploratory per-protocol analyses, HTB was associated with greater improvement in Fatigue, Sleep-related Impairment, and Cognitive Abilities than placebo. Reported side effects were mostly mild, and the proportion of participants reporting side effects did not differ significantly between HTB and placebo.

The findings for Cognitive Abilities in particular warrant further investigation. Although the longitudinal and responder analyses derive from the same instrument and therefore do not represent independent confirmation, both showed effects in the same direction of benefit. Notably, the responder finding remained significant after multiplicity correction, suggesting that perceived cognitive abilities may be an especially useful outcome to prespecify in future HTB trials.

The differing findings for PROMIS Cognitive Function and Cognitive Abilities may partly reflect differences in the constructs emphasized by the two measures. The Cognitive Function short form focuses primarily on perceived difficulties in everyday cognition, including problems with concentration, memory, mental clarity, and information processing, whereas the Cognitive Abilities measure focuses more directly on perceived capacity to perform cognitive tasks, including mental sharpness, memory, concentration, and efficient thinking. In other words, the pattern observed here suggests that HTB may influence the positive or capacity-based aspects of cognition (i.e. cognitive capability) rather than reducing perceptions of cognitive difficulty or impairment. In a community-based, non-clinical population, a measure of perceived cognitive capacity may be more sensitive to modest changes than a measure centered on the occurrence of cognitive difficulties. This interpretation is consistent with the pattern observed here, but the present data cannot establish whether the divergence reflects true construct-specific sensitivity.

Because the per-protocol cohort was restricted to 125 highly adherent participants, the observed improvements may suggest that benefits of HTB depend on consistent daily consumption. Notably, a small double-blind crossover trial in healthy women also reported an early reduction in self-reported fatigue with Tartary Buckwheat cookies.^49^ However, given the smaller sample, the post-randomization definition of this cohort, and the possibility that highly adherent participants differ systematically from others, the present findings should be viewed as exploratory. Nonetheless, they provide additional support for evaluating cognitive abilities and fatigue– and sleep-related outcomes in future trials.

The faster reaction times on the Flanker and Stroop tasks, without corresponding differences in accuracy, suggest faster general processing speed without evidence of compromised task performance. The consistency of this finding across two independent cognitive tasks, together with the observed improvement in self-reported PROMIS Cognitive Abilities, provides convergent, albeit preliminary, evidence of a benefit to cognitive performance. In contrast, no treatment effects were observed for Flanker or Stroop interference reaction time, suggesting that the observed effects were more consistent with general processing speed than cognitive control. Because these computerized outcomes were exploratory findings, they should be interpreted cautiously and warrant prospective confirmation in appropriately powered trials. It is also important to note that the significant treatment × week interaction for Flanker reaction time suggests that the magnitude of the treatment difference varied over time, raising the possibility of an early cognitive benefit. This temporal pattern warrants further investigation to determine whether the cognitive effects of HTB are more pronounced during initial supplementation.

Exploratory epigenetic analyses identified several potentially informative biological signals, although none of the four prespecified epigenetic clocks demonstrated significant between-group differences. Three measures exhibited moderate-to-large relative decreases in the HTB arm: OmniAge.PipekEN.DeltaAge, estimated activated NK-cell fraction, and age-adjusted mitotic age (epiTOC2). These findings may suggest changes in epigenetic age-related measures and immune-cell dynamics. Given the small sample size and absence of findings surviving multiple-testing correction, these results are best viewed as hypothesis-generating. Together with the cognitive findings, they provide a rationale for future dose-response trials incorporating subjective and objective cognitive assessments, including speed–accuracy analyses, alongside targeted biomarkers. Given the known correlations between peripheral inflammatory activation and cognition and mood,^50^ future studies could include epigenetic measures (OmniAge.PipekEN.DeltaAge, epiTOC2, DunedinPACE) as well as markers of immune function and inflammation (NK-cell phenotyping, hs-CRP, IL-6). Ceramide kinase activity, the most strongly enriched pathway in the earlier pilot study,^26^ was not examined in the present analyses but has been implicated in Alzheimer’s disease and may warrant targeted investigation.^51^

The present findings are broadly consistent with the emerging flavonoid literature.^7^ Although this trial used the intervention duration in which flavonoid benefits have been most apparent, it was powered to detect an effect of approximately d = 0.30, roughly twice the pooled estimate of g = 0.15, and may therefore have been underpowered for effects of that size. Prior studies, including the tart cherry trial noted above, have also varied substantially in food matrix, polyphenol composition, dose, population, and cognitive outcomes, and have largely focused on objective cognitive performance. The present study extends this literature by examining the effects of a polyphenol-rich whole food on perceived everyday cognitive function. Given an ongoing need for cost-effective tools to improve and protect cognitive health, the data generated from this trial may provide a useful framework for evaluating whether a high-phytochemical food could serve as a targeted brain health intervention for specific populations.

### Strengths and Limitations

Strengths of this trial include random assignment, double-blind placebo control, a prespecified primary outcome, trial registration, validated patient-reported measures, and complementary computerized and biomarker measures. Several limitations should be noted. Outcomes were self-reported and may be influenced by expectancy and repeated self-monitoring. Recruitment was direct-to-consumer, and participants were self-selected and predominantly White (77%), which limits generalizability. About 22% of randomized participants did not complete a postbaseline survey, and the responder analysis was restricted to the 296 participants with final-week data, so attrition may bias these estimates. The per-protocol and computerized-testing analyses were exploratory comparisons without multiplicity adjustments, and the per-protocol cohort was small. Adherence was also self-reported.

## Conclusions

In this decentralized, randomized, placebo-controlled trial, 12 weeks of daily Himalayan Tartary buckwheat sprout powder did not significantly alter the prespecified primary outcome of self-perceived cognitive function relative to placebo. However, a multiplicity-corrected responder finding for perceived cognitive abilities, together with exploratory evidence of faster processing speed on computerized cognitive tasks, supports further investigation of potential cognitive benefits. Exploratory epigenetic analyses also identified hypothesis-generating signals involving epigenetic age deviation, estimated immune-cell composition, and cellular turnover, although none survived correction for multiple testing. Collectively, these findings support larger, adequately powered dose-response trials that integrate validated subjective and objective cognitive assessments with targeted biological measures to clarify the potential cognitive and physiological effects of polyphenol-rich Himalayan Tartary buckwheat.

### Author contributions

K. White: writing – original draft. M. Chau: writing – original draft, formal analysis. M. Roel: formal analysis, writing – review & editing. V. B. Dwaraka: methodology, formal analysis (epigenetic clock quantification, epigenome-wide and pathway analyses), writing – original draft, writing – review & editing. J. Tartar: writing – review & editing. V. Starratt: writing – review & editing, supervision. S. J. Hewlings: conceptualization, methodology, investigation, supervision, writing – review & editing. J. Chen: conceptualization, methodology, investigation, supervision, writing – review & editing. A. Perlmutter: conceptualization, writing – review & editing. All authors approved the final manuscript.

### Funding

This study was funded by Big Bold Health.

### Data availability

De-identified data may be made available from the corresponding author upon reasonable request.

## Acknowledgements

The authors thank the study participants for their time and acknowledge the Radicle Science study operations team for their work on this trial. We thank Big Bold Health for providing the study products.

## AI-Use

During preparation of this manuscript, the authors used Claude Opus 5.5 (Anthropic) and ChatGPT (OpenAI) to assist with editing, clarity, and review; the authors reviewed and take full responsibility for the content.

## Supplementary Material: Epigenetic Sub-study

**Supplementary Table E1.**
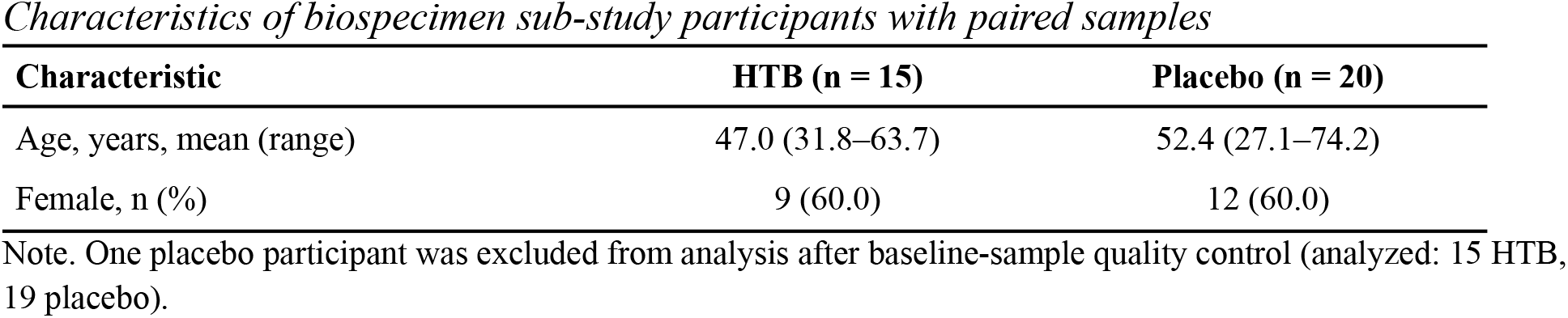
Characteristics of biospecimen sub-study participants with paired samples.

| **Characteristic** | **HTB (n = 15)** | **Placebo (n = 20)** |
|---|---|---|
| Age, years, mean (range) | 47.0 (31.8–63.7) | 52.4 (27.1–74.2) |
| Female, n (%) | 9 (60.0) | 12 (60.0) |

Note. One placebo participant was excluded from analysis after baseline-sample quality control (analyzed: 15 HTB, 19 placebo).

**Supplementary Table E2.** Primary epigenetic clocks: between-arm difference in change from baseline.

| **Clock** | **Cohen's d** | **p** | **FDR** |
|---|---|---|---|
| OMICmAge | 0.40 | .235 | .716 |
| PCGrimAge | 0.26 | .443 | .716 |
| DunedinPACE | 0.16 | .627 | .716 |
| PCPhenoAge | −0.12 | .716 | .716 |

Note. p values are for the treatment × timepoint interaction (age- and sex-adjusted). Cohen's d is the unadjusted between-arm difference in change scores; positive values indicate a larger increase in the HTB arm. FDR = Benjamini–Hochberg false discovery rate across the four clocks.

**Supplementary Table E3.** The three largest between-arm differences among the 9 nominally significant measures (p < .05) in the exploratory screen of 370 clock, mitotic-clock and immune-cell measures.

| **Measure** | **Description** | **Cohen's d** | **p** |
|---|---|---|---|
| OmniAge.PipekEN.DeltaAge | Pipek elastic-net epigenetic age minus chronological age | −0.92 | .009 |
| Activated NK cells | Estimated activated NK-cell fraction (19-cell deconvolution) | −0.89 | .011 |
| tnsc.EAA (epiTOC2) | Estimated cumulative stem-cell divisions, age-adjusted | −0.79 | .023 |

Note. Negative d indicates a smaller increase or larger decrease in the HTB arm than in the placebo arm. No measure remained significant after Benjamini–Hochberg correction across the 370 measures.

**Supplementary Table E4.**
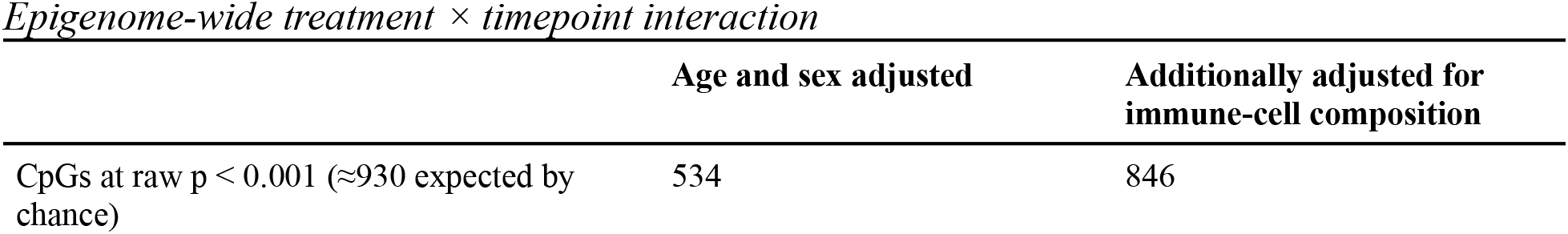
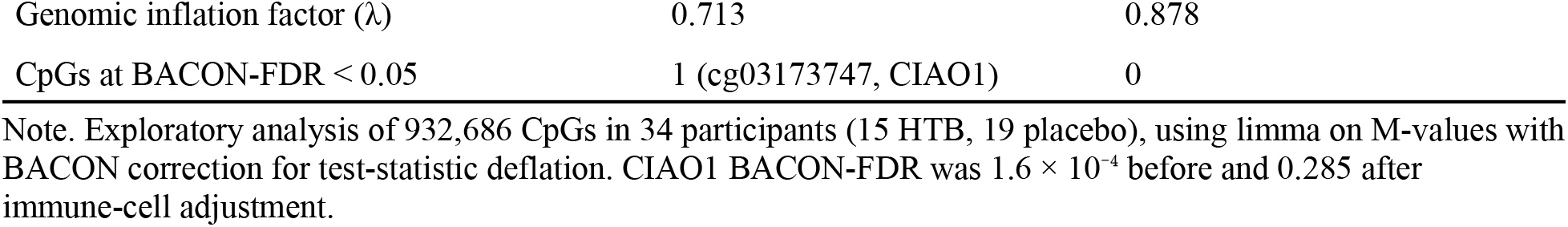
Epigenome-wide treatment × timepoint interaction.

|  | **Age and sex adjusted** | **Additionally adjusted for immune-cell composition** |
|---|---|---|
| CpGs at raw p < 0.001 (≈930 expected by chance) | 534 | 846 |
| Genomic inflation factor (λ) | 0.713 | 0.878 |
| CpGs at BACON-FDR < 0.05 | 1 (cg03173747, CIAO1) | 0 |

Note. Exploratory analysis of 932,686 CpGs in 34 participants (15 HTB, 19 placebo), using limma on M-values with BACON correction for test-statistic deflation. CIAO1 BACON-FDR was 1.6 × 10<sup>−4</sup> before and 0.285 after immune-cell adjustment.

**Supplementary Table E5.** KEGG pathway enrichment among CpGs with raw p < 0.001.

| **Comparison** | **NF-κB signaling** | **TNF signaling** | **Cytokine–receptor interaction** |
|---|---|---|---|
| Treatment × timepoint | 0.066 (4/102) | 0.271 (3/119) | 0.959 (1/292) |
| Within HTB | 0.599 (2/102) | 0.691 (2/119) | 1.000 (0/292) |
| Within placebo | 1.000 (0/102) | 1.000 (0/119) | 0.749 (1/292) |
| Baseline, HTB vs placebo | 0.553 (1/102) | 0.619 (1/119) | 0.822 (1/292) |
| End of intervention, HTB vs placebo | 0.480 (1/102) | 1.000 (0/119) | 0.054 (4/292) |

Note. Values are unadjusted enrichment p-values (overlapping genes/pathway genes) from missMethyl gometh, age and sex adjusted. BH-FDR = 1.0 for every comparison.

**Supplementary Table E6.**
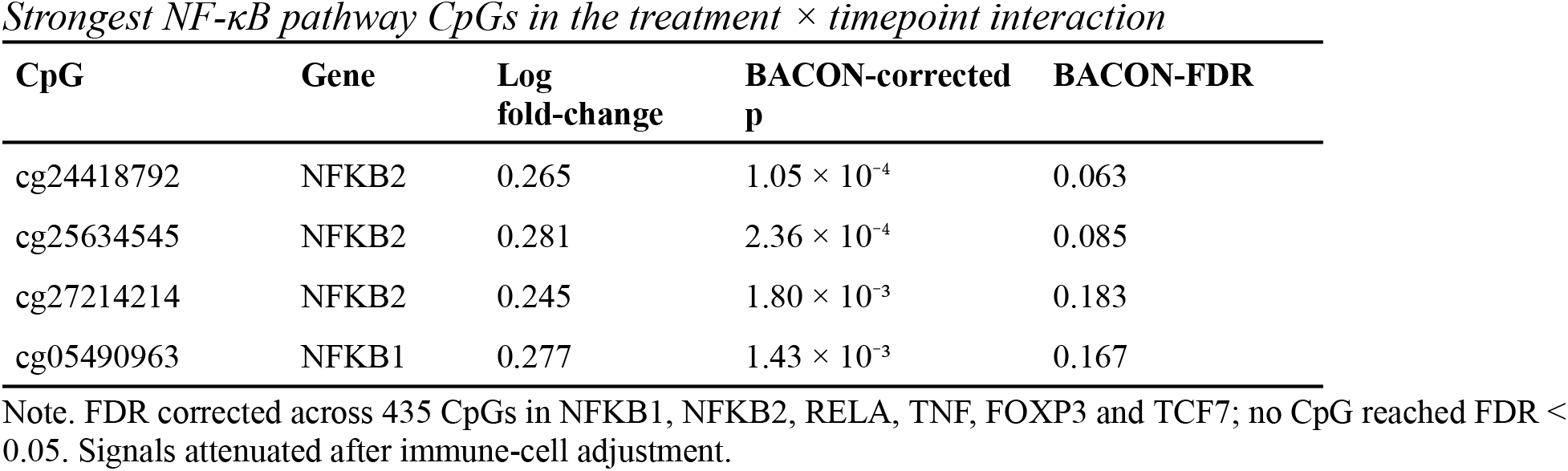
Strongest NF-κB pathway CpGs in the treatment × timepoint interaction.

